# Spinal Cord Injury in Bangladesh (2011–2021): An 11-Year Single-Centre Epidemiological Study

**DOI:** 10.1101/2025.10.08.25337543

**Authors:** Akhlasur Rahman, Md. Shahoriar Ahmed, Farjana Taoheed, Md. Shaikhul Hasan, Sayeed Uddin Helal, Syed Shakawat Hossain, Mohammad Anwar Hossain, Monzurul Alam, Mohammad Sohrab Hossain

## Abstract

Spinal cord injuries (SCI) cause severe, lasting functional impairments and place a significant burden on healthcare systems, especially in developing countries. This retrospective study analyzes epidemiological and demographic data from 3,575 individuals with SCI admitted to the Centre for the Rehabilitation of the Paralysed (CRP) in Bangladesh between 2011 and 2021. Data were collected from patient records, and consent was obtained via phone or in-person. Traumatic SCIs accounted for 92.7% (n=3,155) of cases, primarily caused by falls (56.3%, n=2,014) and road traffic accidents (25.0%, n=892). Most participants were male (85.7%, n=3,065). Paraplegia was the most common condition (58.9%, n=2,104), while tetraplegia was more prevalent in road traffic accident cases (53.7%, n=479). Mortality was 8.7% (n=312), mainly due to pressure sores (39.1%, n=122). Non-traumatic SCIs accounted for 11.8% (n=420), mostly caused by spinal tumors (4.4%, n=157) and transverse myelitis (3.4%, n=121). Among 46 cases of Pott’s disease, females predominated (60.9%, n=28). Geographically, 40.5% (n=1,446) of cases were from the Dhaka division, with Dhaka city reporting the highest number (n=300). These findings offer valuable insights into SCI patterns in Bangladesh and underscore the need for targeted prevention and rehabilitation efforts.

## Introduction

Spinal cord injury (SCI) is a catastrophic neurological condition that results in physical dependence, disability, psychological distress, and financial strain (1). Over the last 30 years, its global incidence has increased from 236 to 1298 cases per million individuals. The estimated worldwide incidence of SCI ranges from 250,000 to 500,000 people annually (2, 3)[3, 11]. In Aisa, the percentage of paraplegic individuals ranges between 18% and 91.97%, while the percentage of quadriplegic individuals ranges from 8.03% to 82% (4, 5). In Bangladesh, the total number of individuals with SCI was 1,035 in 2019 (6)[4].

The consequences of SCI are often devastating, giving rise to numerous complications. These complications can affect nearly all bodily systems, including the cardiorespiratory and genitourinary systems, and may involve localized conditions such as pressure sores or extensive metabolic disorders (1, 7–9). Moreover, the psychosocial impact of SCI can be severe, as patients often face financial burdens, dependence on caregivers, and emotional distress (7).

According to the Global Burden of Disease study, both high-income and developing countries experience high incidence rates of SCI. However, developed nations experienced a lower disease burden due to modernized medical services (10). In contrast, in a low-middle-income country (LMIC) like Bangladesh, both the incidence and burden of disease are significantly higher, leading to severe consequences for survivors and and negatively affecting their socioeconomic status (11). Most SCI survivors are classified under category A of the American Spinal Injury Association Impairment Scale (AIS), accounting for approximately 45% (12). The majority, comprising 59%, have sustained SCI due to traumatic causes (12, 13).

Globally, the leading causeof of SCI are falls and road traffic accidents. Falls from a height, falls with a heavy object and falls on the ground by tripping has been found to be prevalent mechanism for SCI (14). Alarmingly, a study in China revealed that approximately 85.1% of all SCI incidents happened due to falls from high or low height (15). In Bangladesh, the most common cause has been found to be falls from height (42.1%), followed by road traffic accidents (27%) (6).

Although in Bangladesh fall has been the most prevalent cause of SCI, but in recent years, the incidence of road traffic accidents has been rising. This trend raises concern and questions about the underlying contributing factors behind these devastating events (16).

Non-traumatic causes refer to disease-related spinal injuries that result in sensory and motor loss. Certain congenital factors such as spina bifida (17, 18), Chiari malformations (19) and congenital syringomyelia (20) are among the most common types. Inflammatory and autoimmune diseases are also common to cause SCI during their disease prognosis duration (21). Transverse myelitis, acquired demyelinating disorders and neoplasms are widely recognized by healthcare professionals as leading non-traumatic causes of SCI. Degenerative myelopathy (DM) is the most predvalent cause of disease-related spinal cord injury, accounting for 41.1% of all cases, while spinal tumors rank as the second most common cause (6). However, these conditions require further investigation to better understand the underlying mechanisms and influencing factors that may impact the overall rehabilitation outcomes of patients (8, 22). An observational study revealed that the overall number of individuals with spinal cord injury (SCI) in Bangladesh were admitted to non-government institutions (38.65%), whereas only two government hospitals accounted for approximately 30.0% of SCI cases (23). Among these non-government organizations, every year hundreds of SCI survivors seek rehabilitation services from the Centre for the Rehabilitation of the Paralyzed (CRP). (9, 11).

Therefore, the aim of this study is to analyze the trends in the incidence, etiology, and demography of spinal cord injuries over the study period, emphasizing the increasing occurrence of road traffic accidents (RTAs) and falls. In addition, we intend to explore the distribution and risk factors associated with both traumatic and non-traumatic spinal cord injuries. As part of this research, we aim to assess gender-specific disparities in the occurance of Pott’s disease.Finally, we seek to evaluate regional variations in SCI incidence across Bangladesh’s administrative divisions.

## Methods

### Ethical Considerations

The researchers obtained official authorization from the Ethical Review Committee (ERC) of the Center for Rehabilitation of the Paralyzed (CRP) in Savar, Dhaka, Bangladesh, to conduct the study. To collect the necessary information, patients will be contacted via their registered telephone unmbers to obtain consent for the observation and use of data collected during their admission and discharge at CRP.

All information was stored securely in a password-protected database. Unauthorized personnel had no access to the data, thereby ensuring the confidentialty of both individuals and information.

### Data Collection

Records of all patients with SCI from January 2011 to June 2021 were collected from the medical records of the CRP. Socio-demographic data included age, gender, and the number of days spent at CRP. Information on both traumatic and non-traumatic causes of injury was also collected. Traumatic causes included falls from height, road traffic accidents (RTA), falls of heavy objects on the head/neck, falls on the ground, shallow water diving, stab injuries, physical assaults and surgical injuries. For non-traumatic causes, the presence of underlying conditions such as spinal tumors, transverse myelitis, Pott’s disease, cervical myelopathy, spina bifida and neurofibromatosis were investigated. Clinical information included neurological data collected at the time of admission and discharge. The neurological level was assessed according to the American Spinal Injury Association (ASIA) criteria which is considered as the gold standard for assessing and predicting the neurological level and impairment status of a SCI survivors Sensory and motor impairments were assessed, and the level of impairment was determined by the ASIA impairment scale (AIS). All clinical data were recorded at admission and finally at discharge. Recovery from SCI was recorded as either complete or incomplete. Registered patients will be contacted by phone toobtain their consent for the use of data recorded during their admission and discharge at CRP for observational purposes.

### Study setting and participants

To reach the target population, CRP was chosen. CRP is internationally renowned for its SCI department. Each year, CRP provides services to 301 patients, making it one of the largest specialized departments for SCI in the World. This rehabilitation center provides a well-organized acute care and rehabilitation program for patients from across Bangladesh and abroad. During the study period, data were collected from 3,575 patients.

### Analysis

After recording and managing all data within the planned time frame, statistical analyses were conducted using R statistical software. The normality of the data was assessed using the Kolmogorov-Smirnov test and Shapiro-Wilk test with a *p-*value greater than 0.05 indicating normal distribution. Descriptive statistics were performed on socio-demographic data, causative factors and clinical variables. Parametric and continuous data are presented as mean and standard deviation, while discrete and non-parametric data are presented as median with interquartile range and frequency with percentage. To analyze existing correlations, Pearson correlation will be used for continuous data and Spearman rank correlation for ordinal or ranked data.

## Results

### Sociodemographic and Mortality among patients with spinal cord injury

Between 2011 and 2021, we examined the medical records of 3,916 individuals with spinal cord injury (SCI). During the study period, 312 [Figure 1 (STROBE)] individuals passed away from complications related to spinal cord injury (SCI). The primary cause of fatality was pressure sore, accounting for 122 cases (39.10%) We have examined individuals ranging from 5 to 85 years of age. Of the entire population, 3065 (85.73%) were male and 510 (14.27%) were female. So, with a mortality rate of 8.73 per hundred individuals, the final number of SCI patients investigated was 3,575.

**Figure 1.**
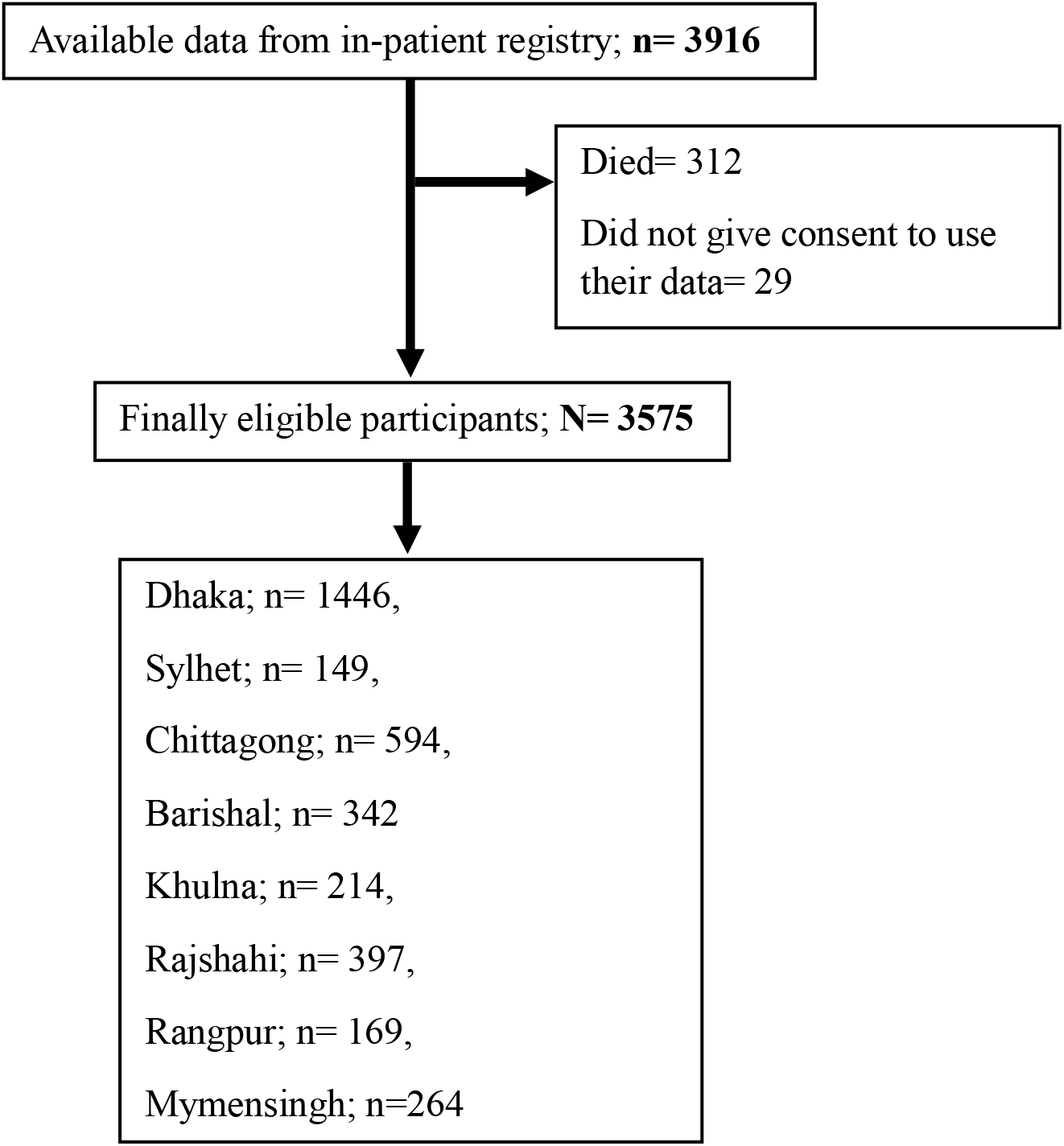
STROBE flowchart

### Clinical and cause-related parameters among survivors of SCI

Table 1 delineates the observed characteristics of SCI survivors. Over fifty percent of the patients are classified as paraplegic (57.76%), including either traumatic (52.13%) or non-traumatic (5.62%) cases. Traumatic spinal cord injury, the predominant cause of SCI in Bangladesh, may result from several incidents.

**Table 1.** Characteristics of SCI survivors attending rehabilitation.

| Demographic Variables | Diagnosis |  |  |  | Total (N=3575) |
| --- | --- | --- | --- | --- | --- |
|  | TP (n= 1772) | TT (n= 1383) | NTP (n= 332) | NTT (n= 88) |  |
| Age (years) |  |  |  |  |  |
| 5-15 | 114(6.43) | 80(5.78) | 24(7.23) | 4(4.55) | 222(6.21) |
| 16-25 | 455(25.68) | 344(24.87) | 89(26.81) | 30(34.09) | 918(25.68) |
| 26-35 | 512(28.89) | 378(27.33) | 105(31.63) | 23(26.14) | 1018(28.48) |
| 36-45 | 332(18.74) | 277(20.03) | 53(15.96) | 17(19.32) | 679(18.99) |
| 46-55 | 224(12.64) | 189(13.67) | 47(14.16) | 9(10.23) | 469(13.12) |
| 56-65 | 107(6.04) | 85(6.15) | 5(1.51) | 2(2.27) | 199(5.57) |
| 66-75 | 27(1.52) | 25(1.81) | 8(2.41) | 2(2.27) | 62(1.73) |
| 76-85 | 1(0.06) | 5(0.36) | 1(0.30) | 1(1.14) | 8(0.22) |
| Gender |  |  |  |  |  |
| Male | 1511(85.27) | 1199(86.70) | 281(84.64) | 74(84.09) | 3065(85.73) |
| Female | 261(14.73) | 184(13.30) | 51(15.36) | 14(15.91) | 510(14.27) |
| AIS category |  |  |  |  |  |
| Complete A | 1053(59.42) | 803(58.06) | 201(60.54) | 54(61.36) | 2111(59.05) |
| Incomplete B | 279(15.74) | 217(15.69) | 47(14.16) | 13(14.77) | 556(15.55) |
| Incomplete C | 226(12.75) | 180(13.02) | 51(15.36) | 14(15.91) | 471(13.17) |
| Incomplete D | 193(10.89) | 167(12.08) | 30(9.04) | 7(7.95) | 397(11.10) |
| Incomplete E | 21(1.19) | 16(1.16) | 3(0.90) | 0(0.00) | 40(1.12) |
| Days stay at CRP |  |  |  |  |  |
| 1-50 | 356(20.09) | 275(19.88) | 75(22.59) | 17(19.32) | 723(20.22) |
| 51-100 | 402(22.69) | 287(20.75) | 67(20.18) | 16(18.18) | 772(21.59) |
| 101-150 | 543(30.64) | 443(32.03) | 96(28.92) | 29(32.95) | 1111(31.08) |
| 151-200 | 276(15.58) | 220(15.91) | 52(15.66) | 17(19.32) | 565(15.80) |
| 201-250 | 103(5.81) | 96(6.94) | 28(8.43) | 5(5.68) | 232(6.49) |
| Above 250 | 92(5.19) | 62(4.48) | 14(4.22) | 4(4.55) | 172(4.81) |

TP, Traumatic Paraplegic; TT, Traumatic Tetraplegic; NTP, Non-Traumatic Paraplegic; NTT, Non-Traumatic Tetraplegic. Data are presented as n (%).

Figure 2 illustrates the frequency distribution of traumatic causes of SCI from 2011 to 2021. The data reveal that fall-related incidents were the most prevalent cause, followed by road traffic accidents (RTAs) in each year. The highest frequency (n=198) of fall related injuries were prevalent in the year 2017. Although the frequency of fall related injuries is declining each year. On the other hand, RTA related SCI are increasing day by day and have been found highest (n=120) in the year 2015. The frequency of RTA related SCI has dropped slightly after 2015, but in recent times the number of SCI caused by traffic accident is rising. The next most prevalent cause of SCI is due to physical assault and has a peak count of 14 cases in 2014. The cases of SCI due to physical assault are also declining year by year. Post surgical complications and shallow water diving cases of SCI patients are close to each other when explored for total count from 2011 to 2021. In case of bull attack related SCI, 25 cases were found in 2014. This count for bull attack has also declined rapidly in recent years. Violent attacks such as stabbing and gunshot were also found responsible for SCI and the total counts of cases after eleven years are 36 and 8, respectively. The least found cases within the observed year is due to sports injury and counts for only 2 cases.

**Figure 2.**
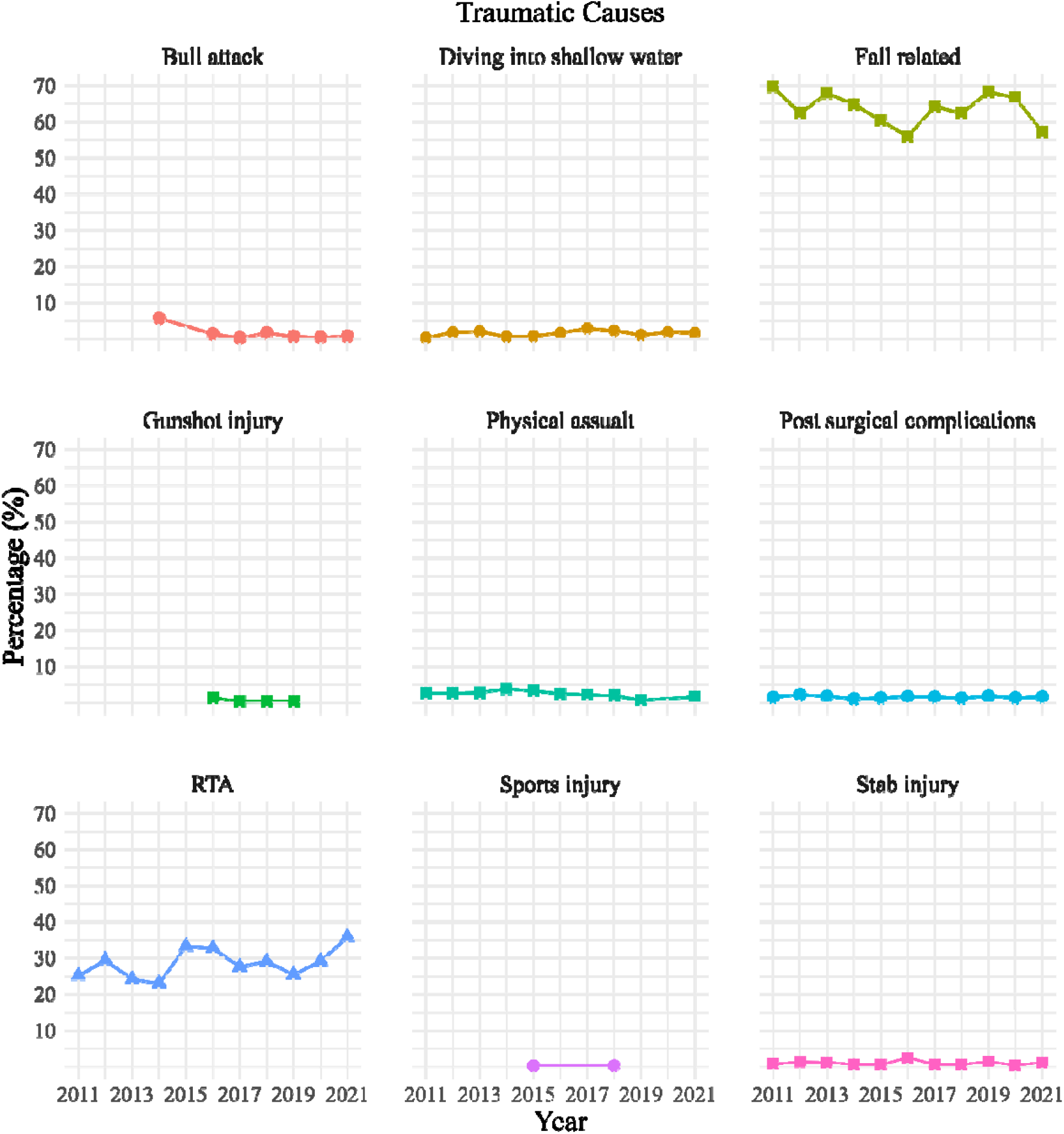
Frequency distribution of traumatic causes of SCI. The x-axis holds the years from 2011 to 2021, and the y-axis is the percentages of occurrences of different traumatic incidents.

After exploring the traumatic causes of SCI, it is crucial to illustrate the most rising cause of SCI. To outline the type of paralysis, Figure 3 has been plotted to understand the outcome following an RTA. It is visible that the majority of SCI cases after RTA are tetraplegic (n= 527) out of 949 total cases found within observed years. In 2015, the count of RTA-related SCI was 120 and the majority of them were tetraplegic. Although the count declined for some years the count of RTA-related SCI is rising rapidly and the worst-case scenario is that, majority of them have been found to be diagnosed as tetraplegic (n=86) in 2022.

**Figure 3.**
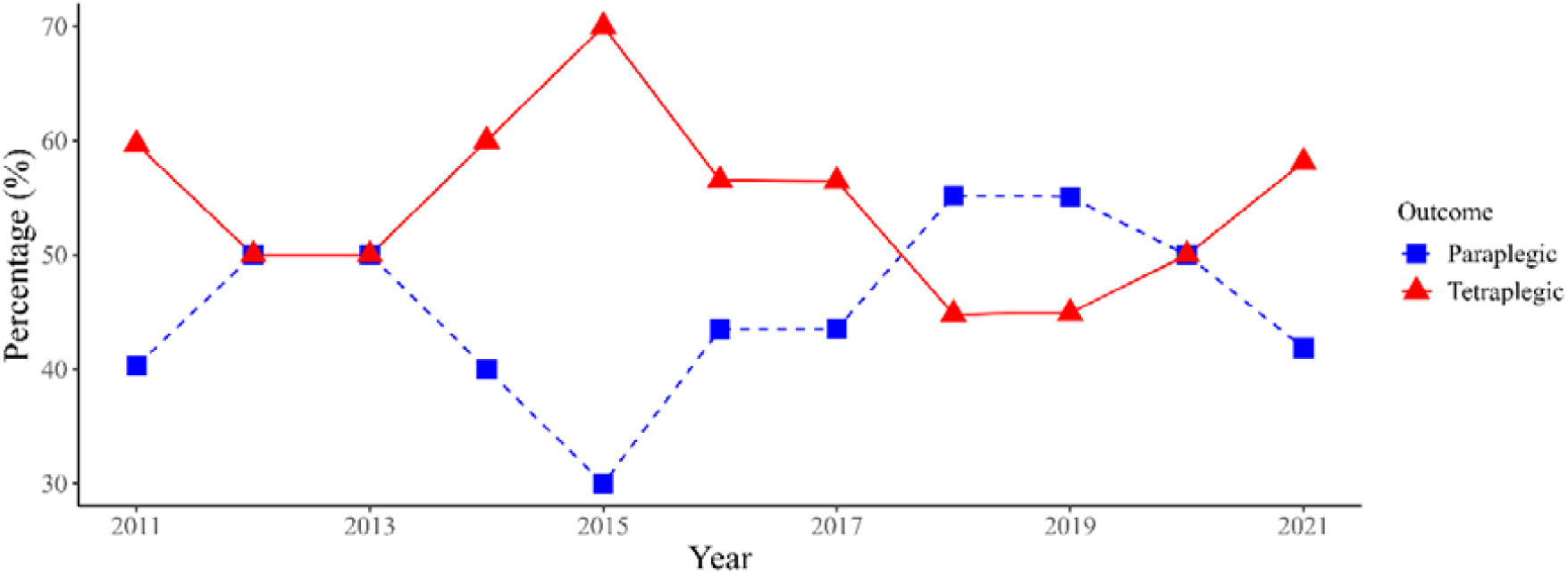
Frequency distribution of road traffic accident. The x-axis holds the years from 2011 to 2021, and the y-axis is the percentages of occurrences of road traffic accident.

After observing eleven years of data, non-traumatic SCI cases were found 420. Non-traumatic causes of SCI have been illustrated in **Figure 4**. The predominant non-traumatic cause of SCI was identified as spinal tumour (n=157), followed by transverse myelitis (n=121). The cases due to idiopathic pathological cause (n=47) and Pott’s disease (n=46) were also prevalent. In 34 non-traumatic cases, cervical myelopathy was found to be the etiology of SCI. Other rare non-traumatic causes of SCI included spina bifida (n=8), Neurofibromatosis (n=6) and autoimmune disease (n=1). Among all those pathological cases, Pott’s disease is known as spinal tuberculosis (TB), and if observed closely, we might observe some gender predominance.

**Figure 4.**
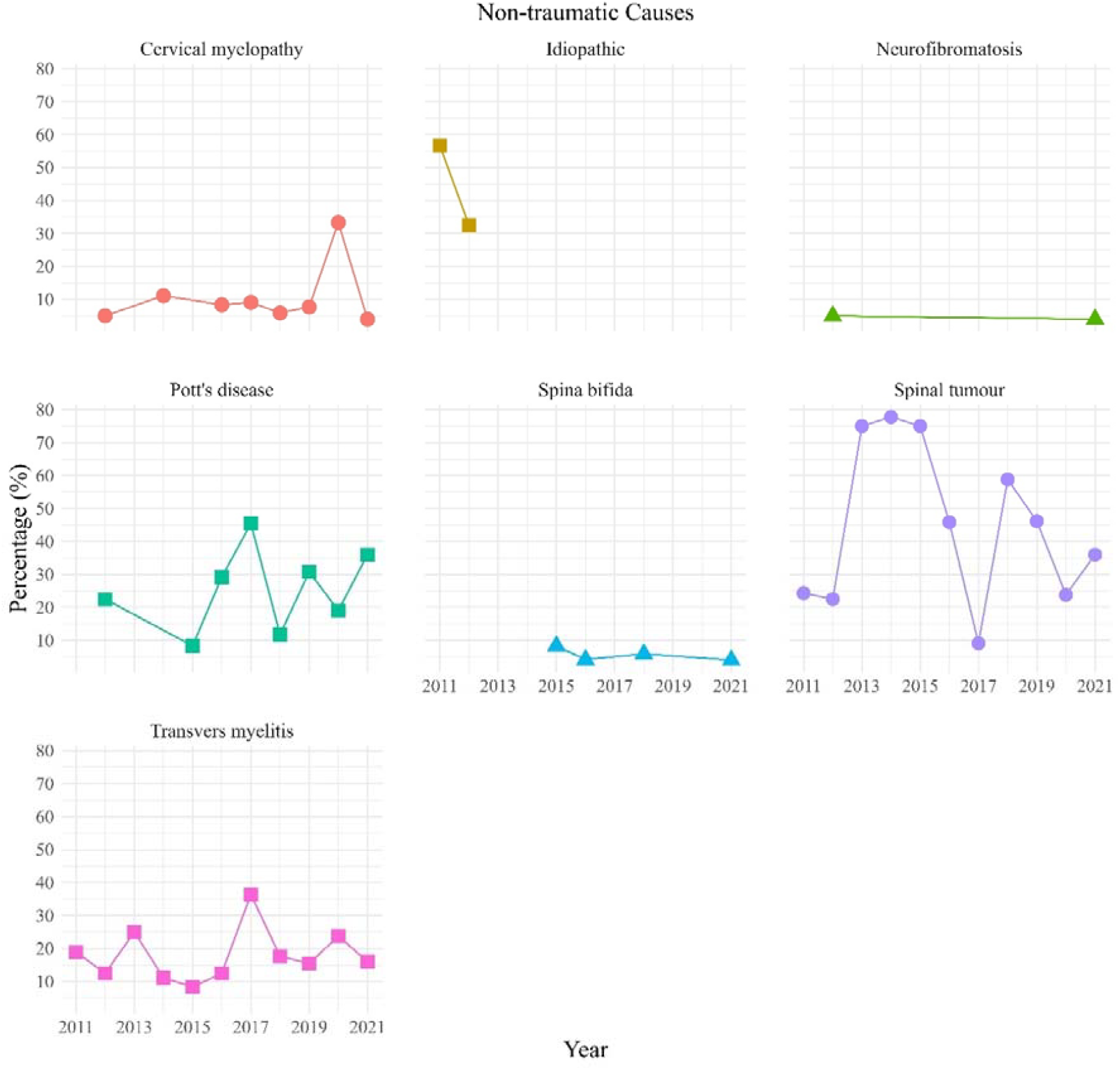
Frequency distribution of non-traumatic causes of SCI. The x-axis holds years from 2011 to 2021, and y-axis holds percentage of different non-traumatic causes.

Figure 5 illustrates the distribution seen in the prevalence of Pott’s illness between genders. It is evident that women in Bangladesh are more susceptible to Pott’s illness, with a notable rise in its incidence in recent years.

**Figure 5.**
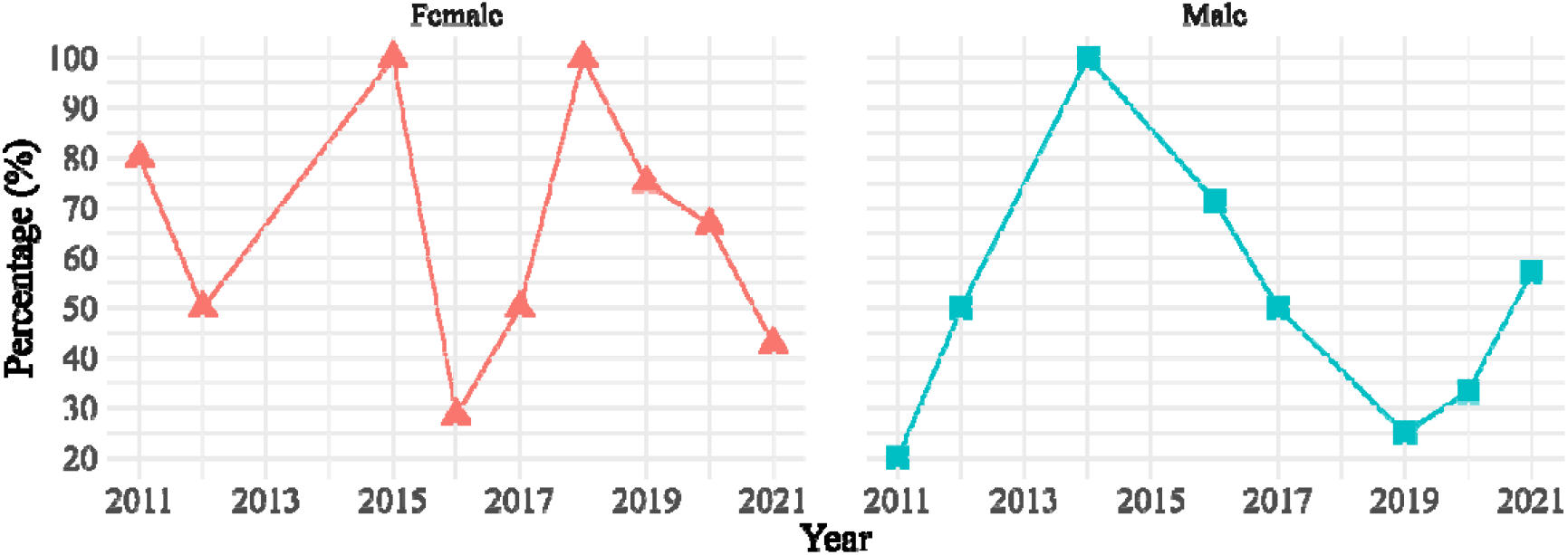
Frequency distribution of Pott’s disease according to gender. The x-axis holds years from 2011 to 2021, and y-axis holds percentage of occurrence among both genders year.

### Geographical distribution across the administrative divisions of the country

We have observed data coming from different branches of CRP containing yearly incidence rates from 2011 to 2021. **Figure 6** depicts the incidence distribution of SCI among the eight divisions of Bangladesh. Dhaka has been found to be the focal point, with 1446 patients, accounting for 39.94% of total instances. The second highest incident of SCI took place in Chittagong with a total case of 594 patients, covering 16.61% of total SCI incidents. Rajshahi has the third place for the most counted SCI incidents numbering for 397 cases, rounding up 11.10% total cases. The closest count was found in Barishal (342; 9.56%) when compared with Rajshahi. Mymenshingh (264; 7.40%) and Khulna (214; 5.98%) rank for fifth and sixth division in terms of SCI incidences within the explored years. Divisions like Rangpur (3.78%) and Sylhet (2.50%) had far lower rates in contrast to other divisions of Bangladesh. The map of Dhaka division shows the hotspot district/zilla for SCI. The highest incidence in Dhaka division was found for Dhaka city (n=300). Faridpur was the second highest incidence spot with a total number of 248 SCI patients. The remaining districts are ranked as follows: Gazipur (n=222), Gopalganj (n=167), Kishoreganj (n=143), Madaripur (n=130), Manikganj (n=52), Munshiganj (n=48), Narayanganj (n=42), Narshingdi (n=37), Rajbari (n=32), Shariatpur (n=13), Tangail (n=12). Both maps employ color gradients to illustrate the numerical intensity of SCI occurrences, emphasizing notable divisional discrepancies.

**Figure 6.**
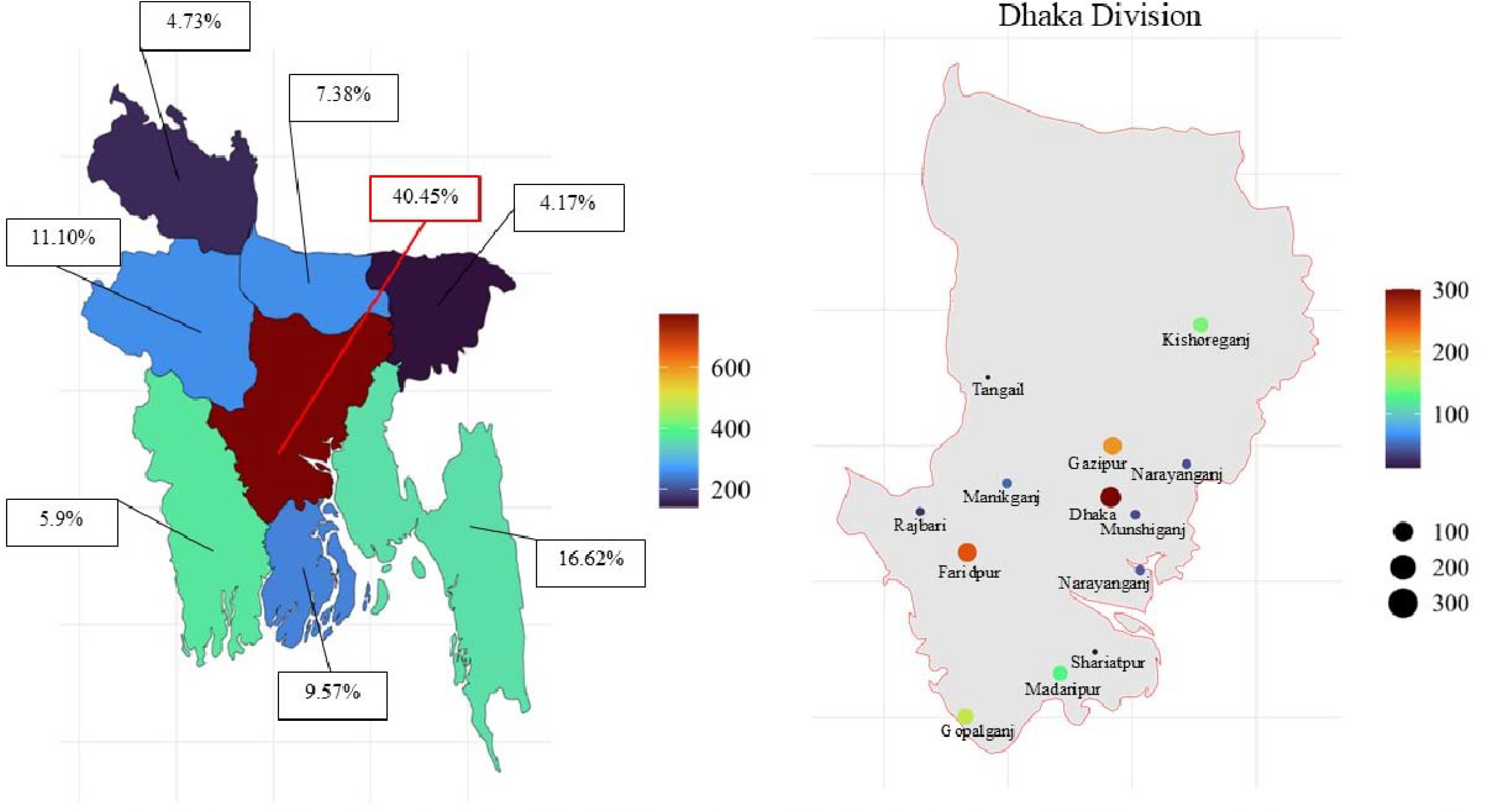
Distribution of data throughout the eiglliitdivisions of Bangladesh. The designated divisions are: Dhaka, 40.45%; Sylhet, 4.17%; Chittagong, 16.61%; Barishal, 9.56%; Khulna, 5.98%; Rajshahi, 11.10%; Rangpur, 4.73%; Mymensingh, 7.40%. Tlhe color gradients signify a munerical intensity of SCI incidence across divisions. The Dhaka division has been extensively examined to identify the hotspot regions of SCI incidence.

## Discussion

In our study, we summarized the general characteristics of the SCI population based on socio-demographic, clinical and geographic parameters. Most SCI cases occurred among individuals aged 16 to 55 years. Men were more susceptible to traumatic causes of SCI than women. The most common traumatic causes were falls (56.34%) and RTA (24.95%). Alarmingly, the incidence of RTAs has been increasing in recent years. The incidence of RTA might be reduced if the location and time of occurrence are investigated. Additionally, data-driven interventions, such as awareness campaigns, infrastructure improvements, traffic law enforcement, annual driver’s license renewals, and collaboration with legislators to fortify governance, can significantly improve road safety and injury prevention.

In case of non-traumatic SCI causes, the most common disease condition that leadingto SCI is spinal tumor. According to the literature, both spinal tumor and transverse myelitis predominantly affect the thoracic region (24, 25). Recent evidence suggests that approximately 76.5% of patients with spinal tumors undergo surgery and postoperative complications such as neurological impairment arise in about 61.3% of these cases (24). The second most common disease-related cause of SCI is transverse myelitisan inflammatory disorder of the spinal cord that leads to neurological deficits.The final outcome often involves a clearly defined neurological level, tipically measured using the ASIA impairment scale (25). An important finding from this study concerns Pott’s disease. Pott’s disease or spinal tuberculosis (TB) is caused by infection with *Mycobacterium tuberculosis* and is categorized as extra-pulmonary TB affecting the spinal column (26). Delayed diagnosis and treatment can result severe outcomes, including permanent spinal cord injury (27). Despite widespread awareness of TB in Bangladesh (28), the number of TB-related SCI is rising, with a significantly higher occurrence among women. This highlights the critical need for increasing awareness, addressing key social determinants of health, ensuring equitable access to affordable healthcare, inclusive education, and social support particularly for women. Additionally, disseminating general information about Pott’s disease among Bangladeshi people is essential for early detection and prevention.

The findings also reveal that the Dhaka division accounts for the highest proportion of SCI cases, at 40.45%, identifying it as a notable hotspot. This concentration may be attributed to several factors, including high traffic density and intense movement of people at a certain time. This indicates the necessity to search for the causes of accidents and co-founding factors related to these accidents. Existing literature strongly suggests that inadequate traffic signals, insufficient traffic lights at night, unskilled driving and unsafe overtaking maneuvers contribute to traffic accidents (29). In contrast, the Barishal and Mymensingh divisions had markedly low rates, subsequently 9.57% and 7.38%. Uddin et al. (2021) conducted an observational study to explore the clinical parameters along the geographic distribution of SCI occurrence across Bangladesh. In our study, we have summarized the SCI occurrence up to level 2 administrative areas that may lead us to observe the hotspot of SCI in Bangladesh. The disproportionate incidence of SCI in some areas, particularly in Dhaka division, requires targeted investigation to address the causes of high incidence, including the implementation of awareness initiatives. This suggests further research to find causes of high incidence in Dhaka in comparison to other divisions.

We hope that this long-term epidemiological research from Bangladesh will advance our knowledge of spinal cord injury (SCI) in low-resource settings. The study will highlight the concerning growth in road traffic accident (RTA)-related SCI and chronic fall-related injuries over twelve years. Breaking down etiological patterns highlights context-specific risks, such as the disproportionate burden of traumatic SCI among men in high-traffic urban areas like Dhaka and the gender disparity in non-traumatic SCI, particularly the suspected female predominance in Pott’s disease, which will highlight neglected spinal tuberculosis management. Geographical mapping of SCI hotspots exposes regional inequalities, with Dhaka’s high prevalence of traffic congestion and hazardous working conditions. The study recommends targeted road safety measures, strengthening workplace regulations to minimize fall risks, and public health campaigns to prevent delayed diagnosis of illnesses like Pott’s disease. Based on clinical, demographic, and spatial perspectives, this research advances the global SCI literature and provides policymakers and healthcare providers with evidence to design context-specific interventions to reduce the socioeconomic and personal impact of SCI in Bangladesh and similar low-resource settings.

## Conclusion

The epidemiology of spinal cord injury (SCI) in Bangladesh has been studied in this eleven-year study, which identifies traumatic causes primarily falls and road traffic accidents as the main causes. Notable findings include the gender disparity in non-traumatic cases, especially among females affected by Pott’s disease, and a rising prevalence of SCI related to road traffic accidents (RTAs). Dhaka, due to its high traffic congestion and other causative factors, has emerged as a hotspot for SCI. These findings highlighted the need for targeted interventions, including fall prevention strategies, improved road safety measures, strengthened occupational regulations, and increase public awareness. These observations can help healthcare providers and policymakers in developing context-specific strategies to minimize the incidence of SCI in Bangladesh.

## Data Availability

All data produced in the present study are available upon reasonable request to the authors

## Acknowledgments

The authors sincerely thank the Centre for the Rehabilitation of the Paralysed (CRP), Savar, Dhaka, Bangladesh, for granting access to medical records and supporting data collection. We would like to express our gratitude to Yeasin, Jahid, Rubayet and Hamim for their valuable assistance in data collection and follow-up activities. Special thanks also go to the medical record officers, research assistants, and clinical staff of CRP for their continuous support throughout this study.

## Funding Statement

This research received no specific grant from any funding agency in the public, commercial, or not-for-profit sectors.

## Declaration of Interest Statement

The authors declare no potential conflicts of interest with respect to the research, authorship, and/or publication of this article.

## Notes

### Competing Interest Statement

The authors have declared no competing interest.

### Funding Statement

This study did not receive any funding

### Author Declarations

Ethical Review Committee (ERC) of the Center for Rehabilitation of the Paralyzed (CRP) gave ethical approval for this work.

## References

1. Anjum A, Yazid MDi, Fauzi Daud M, Idris J, Ng AMH, Selvi Naicker A, et al. Spinal cord injury: pathophysiology, multimolecular interactions, and underlying recovery mechanisms. International journal of molecular sciences. 2020;21(20):7533.

2. Khorasanizadeh M, Yousefifard M, Eskian M, Lu Y, Chalangari M, Harrop JS, et al. Neurological recovery following traumatic spinal cord injury: a systematic review and meta-analysis. Journal of Neurosurgery: Spine. 2019;30(5):683–99.

3. Singh A, Tetreault L, Kalsi-Ryan S, Nouri A, Fehlings MG. Global prevalence and incidence of traumatic spinal cord injury. Clinical epidemiology. 2014:309–31.

4. Ning G-Z, Wu Q, Li Y-L, Feng S-Q. Epidemiology of traumatic spinal cord injury in Asia: a systematic review. The journal of spinal cord medicine. 2012;35(4):229–39.

5. Eli I, Lerner DP, Ghogawala Z. Acute traumatic spinal cord injury. Neurologic clinics. 2021;39(2):471–88.

6. Uddin T, Islam MT, Hossain M, Hossain MS, Salek A, Islam MJ, et al. Demographic and Clinical Characteristics of Persons With Spinal Cord Injury in Bangladesh: Database for the International Spinal Cord Injury Community Survey 2023. Neurotrauma Reports. 2023;4(1):598–604.

7. New PW. Secondary conditions in a community sample of people with spinal cord damage. The journal of spinal cord medicine. 2016;39(6):665–70.

8. Rahman H. Social experience of men with paraplegic spinal cord injury in BANGLADESH: Bangladesh Health Professions Institute, Faculty of Medicine, the University …; 2016.

9. Quadir MM, Sen K, Sultana MR, Ahmed MS, Taoheed F, Andalib A, et al. Demography, diagnosis and complications of spinal cord injury patients in a rehabilitation center of Bangladesh. 2023.

10. Ding W, Hu S, Wang P, Kang H, Peng R, Dong Y, et al. Spinal cord injury: the global incidence, prevalence, and disability from the global burden of disease study 2019. Spine. 2022;47(21):1532–40.

11. Hossain MS, Islam MS, Rahman MA, Glinsky JV, Herbert RD, Ducharme S, et al. Health status, quality of life and socioeconomic situation of people with spinal cord injuries six years after discharge from a hospital in Bangladesh. Spinal Cord. 2019;57(8):652–61.

12. Hachem LD, Ahuja CS, Fehlings MG. Assessment and management of acute spinal cord injury: From point of injury to rehabilitation. The journal of spinal cord medicine. 2017;40(6):665–75.

13. Hossain M, Rahman M, Bowden JL, Quadir M, Herbert R, Harvey L. Psychological and socioeconomic status, complications and quality of life in people with spinal cord injuries after discharge from hospital in Bangladesh: a cohort study. Spinal Cord. 2016;54(6):483–9.

14. Zhang Z-R, Wu Y, Wang F-Y, Wang W-J. Traumatic spinal cord injury caused by low falls and high falls: a comparative study. Journal of orthopaedic surgery and research. 2021;16(1):222.

15. Wang Z-M, Zou P, Yang J-S, Liu T-T, Song L-L, Lu Y, et al. Epidemiological characteristics of spinal cord injury in Northwest China: a single hospital-based study. Journal of orthopaedic surgery and research. 2020;15(1):214.

16. Alam ABMK. Road traffic accidents in Bangladesh. Journal of Bangladesh College of Physicians & Surgeons. 2018;36(4):137.

17. New PW, Eriks-Hoogland I, Scivoletto G, Reeves RK, Townson A, Marshall R, et al. Important clinical rehabilitation principles unique to people with non-traumatic spinal cord dysfunction. Topics in spinal cord injury rehabilitation. 2017;23(4):299–312.

18. Hopson B, Rocque BG, Joseph DB, Powell D, McLain AB, Davis RD, et al. The development of a lifetime care model in comprehensive spina bifida care. Journal of pediatric rehabilitation medicine. 2018;11(4):323–34.

19. New PW, Marshall R. International Spinal Cord Injury Data Sets for non-traumatic spinal cord injury. Spinal cord. 2014;52(2):123–32.

20. Nakamura M, Ishii K, Watanabe K, Tsuji T, Matsumoto M, Toyama Y, et al. Clinical significance and prognosis of idiopathic syringomyelia. Clinical Spine Surgery. 2009;22(5):372–5.

21. Molinares DM, Gater DR, Daniel S, Pontee NL. Nontraumatic spinal cord injury: epidemiology, etiology and management. Journal of personalized medicine. 2022;12(11):1872.

22. Kisala PA, Tulsky DS, Choi SW, Kirshblum SC. Development and psychometric characteristics of the SCI-QOL Pressure Ulcers scale and short form. The Journal of Spinal Cord Medicine. 2015;38(3):303–14.

23. Kumar R, Lim J, Mekary RA, Rattani A, Dewan MC, Sharif SY, et al. Traumatic spinal injury: global epidemiology and worldwide volume. World neurosurgery. 2018;113:e345–e63.

24. Ge L, Arul K, Mesfin A. Spinal cord injury from spinal tumors: prevalence, management, and outcomes. World Neurosurgery. 2019;122:e1551–e6.

25. Lim PA. Transverse myelitis. Essentials of physical medicine and rehabilitation. 2019:952.

26. Glassman I, Nguyen KH, Giess J, Alcantara C, Booth M, Venketaraman V. Pathogenesis, diagnostic challenges, and risk factors of Pott’s disease. Clinics and Practice. 2023;13(1):155–65.

27. Kamara E, Mehta S, Brust JC, Jain AK. Effect of delayed diagnosis on severity of Pott’s disease. International orthopaedics. 2012;36(2):245–54.

28. Masud Rana MR, Abu Sayem AS, Reazul Karim RK, Nurul Islam NI, Rafiqul Islam RI, Tunku Kamarul Zaman TKZ, et al. Assessment of knowledge regarding tuberculosis among non-medical university students in Bangladesh: a cross-sectional study. 2015.

29. WHO V. Global status report on road safety 2018. World Health Organization. 2018.

